# An extended SEIARD model for COVID-19 vaccination in Mexico: analysis and forecast

**DOI:** 10.1101/2021.04.06.21255039

**Authors:** Ángel G. C. Pérez, David A. Oluyori

**Affiliations:** Facultad de Matemáticas, Universidad Autónoma de Yucatán, Mérida, Yucatán, Mexico; Department of Mathematics, School of Physical Science, Ahmadu Bello University, Zaria, Kaduna State, Nigeria

## Abstract

In this study, we propose and analyse an extended SEIARD model with vaccination. We compute the control reproduction number ℛ_*c*_ of our model and study the stability of equilibria. We show that the set of disease-free equilibria is locally asymptotically stable when ℛ_*c*_ < 1 and unstable when ℛ_*c*_ > 1, and we provide a sufficient condition for its global stability. Furthermore, we perform numerical simulations using the reported data of COVID-19 infections and vaccination in Mexico to study the impact of different vaccination, transmission and efficacy rates on the dynamics of the disease.

## 1 Introduction

The Coronavirus Disease 2019 (COVID-19) pandemic, caused by the Severe Acute Respiratory Syndrome Coronavirus 2 (SARS-CoV-2) has caused a worldwide crisis due to its effects on society and global economy. Due to the absence of specific anti-COVID-19 medical treatments, most countries had been relying on non-pharmaceutical interventions, such as wearing of face masks, social/physical distancing, partial/total lockdown, travel restrictions, and closure of schools and work centres, in order to curtail the spread of the disease before December 2020. However, these measures have been insufficient to mitigate the pandemic globally as medical facilities were overstretched and death toll heightened.

Vaccination has been an effective strategy in combating the spread of infectious diseases, e.g., pertussis, measles, and influenza. Historically, the eradication of smallpox has been considered as the most remarkable success of vaccination ever recorded [1]. So far, the development and testing of vaccines against SARS-CoV-2 has occurred at an unprecedented speed and, in the last months, several vaccines have been approved for use in many countries, and their deployment is already underway. In a pandemic situation such as this, current preventive vaccines consisting of inactivated viruses do not protect all vaccine recipients equally as the protection conferred by the vaccine is dependent on the immune status of the recipient [2].

Over the past few decades, a large number of simple compartmental mathematical models with vaccinated population have been proposed in the literature to assess the effectiveness of vaccines in combatting the infectious diseases [3–12]. With the recent development of anti-COVID vaccines, several models have been proposed to provide insight into the effect that inoculation of a certain portion of the population will have on the dynamics of the COVID-19 pandemic. The authors in [13] studied an SEIHRDV model and an SMEIHRDV model (the latter including a semi-susceptible class) and fitted the parameters using data from several countries to evaluate the effect of social distancing and vaccination on controlling COVID-19. Another model was studied in [14] to compare the outcomes of single-dose and two-dose anti-COVID vaccination regimes. The global stability of a two-strain COVID-19 model with vaccination against one strain was studied in [15]. In [16], the effect of immunity, vaccination, and reinfection with changing parameters was analysed using an SEVIS model, while in [17] an SIQRD model was used to simulate several scenarios of vaccine delivery in Indonesia.

Since it is known that individuals infected with SARS-CoV-2 can transmit the virus to other people without presenting symptoms of the disease, some authors have proposed models that distinguish between symptomatic and asymptomatic infections. For instance, Moore *et al*. [18] proposed an age- and region-structured model to simulate the rollout of a two-dose vaccination programme in the UK using the Pfizer–BioNTech and Oxford–AstraZeneca vaccines. Vilches *et al*. [19] used an agent-based transmission model to project the impact of a two-dose vaccination campaign with the Pfizer–BioNTech and Moderna vaccines in Ontario, while Olivares & Staffetti [20] conducted a sensitivity analysis and uncertainty quantification of an SEIsIaQR model with vaccination strategy. A three-patch metapopulation epidemic model structured by risk was used to investigate several vaccination scenarios in Mexico City in [21]. Some other COVID-19 models with vaccination can be found in [22–26].

In mid-November 2020, Mexico passed the mark of 1,000,000 confirmed cases and 100,000 deaths due to COVID-19. On 11 December 2020, the Mexican government’s medical safety commission approved the emergency use of the Pfizer–BioNTech coronavirus vaccine, with the first 250,000 doses intended for health workers. The inoculation of frontline health personnel started that year on 24 December. Four other COVID-19 vaccines were approved between January and February 2021, and vaccination of people over 60 years of age with the AstraZeneca vaccine began in February. The first vaccines to be applied in Mexico followed a two-dose regime, until the single-dose CanSino vaccine began to be applied to a portion of the population in April 2021. The Johnson & Johnson vaccine, also single-dose, was deployed in June 2021 to inoculate the population over age 18 in the municipalities of the northern border. The government expects to have vaccinated all adult population with at least one dose by October 2021.

In this paper, we propose a differential equation model to simulate the application of a two-dose vaccine against COVID-19, considering the possibility of vaccine leakiness and asymptomatic infections. The motivation of this study is derived from the work of the authors in [27, 28], who considered an SEIARD mathematical model to investigate the outbreak of COVID-19 in Mexico. Therefore, in the present work, we incorporate the vaccination component to the model in [28] to derive an extended SEIARD model to examine the effectiveness of the COVID-19 jabs which are currently being deployed to many countries to help combat the raging pandemic situation.

The rest of this paper is organized as follows. In Section 2, we present the equations and assumptions of the extended SEIARD model with vaccination. In Section 3, we perform a theoretical analysis of the model, compute its control reproduction number and study the stability of the disease-free equilibria. In Section 4, we carry out numerical simulations using reported data on COVID-19 infections and vaccination in Mexico. Lastly, we provide some discussions and concluding remarks in Section 5.

## 2 Model formulation

To derive the mathematical model, we subdivide the unvaccinated population into susceptible (*S*), exposed (*E*), symptomatic infectious (*I*), asymptomatic infectious (*A*), and recovered (*R*). The number of individuals in each subpopulation at time *t* is denoted by *S*(*t*), *E*(*t*), etc. Susceptible individuals become exposed by contact with symptomatic infectious individuals at a rate *β*_1_ and by contact with asymptomatic infectious individuals at a rate *β*_2_. The exposed individuals become infectious at a rate *w*: a proportion *p*_1_ of them will show symptoms of the disease, while the rest remains asymptomatic. We assume that the symptomatic class has a disease-induced death rate, denoted by *δ*_1_ (our model does not consider deaths not related to COVID-19). Both symptomatic and asymptomatic infectious people recover at a rate *γ*.

We also assume that the susceptible population *S* is vaccinated at a rate *v* ≥ 0 (the number of first doses administered per day at time *t* is given by *vS*(*t*)). Individuals who have received only the first dose of the vaccine are included in the class *V*_1_, and they move to the class *V*_2_ upon receiving the second dose, which occurs at a rate *θ*. Both *V*_1_ and *V*_2_ are considered susceptible. Due to vaccine leakiness, vaccination of an individual does not completely remove the risk of infection. Hence, we also assume that the vaccinated population can become exposed (*E*_*V*_), symptomatic infectious (*I*_*V*_) and asymptomatic infectious (*A*_*V*_). Individuals in the class *V*_1_ (respectively, *V*_2_) move to the class *E*_*V*_ due to contact with symptomatic infectious people at a rate (1−*η*_1_)*β*_1_ (respectively, (1 − *η*_2_)*β*_1_) and by contact with the asymptomatic infectious at a rate (1 − *η*_1_)*β*_2_ (respectively, (1 − *η*_2_)*β*_2_), where *η*_1_ is the efficacy of the vaccine after one dose (*η*_2_ is the efficacy of the vaccine after two doses). The infectivity of individuals in the *I*_*V*_ and *A*_*V*_ classes is reduced by a factor 1 − *q* with respect to that of individuals in the *I* and *A* classes, where *q* ∈ [0, 1].

The population in the class *E*_*V*_ becomes infectious at a rate *w*; we assume that the proportion of people from this class who become symptomatic infectious is *p*_2_, which may be different from that of unvaccinated people due to the effect of the vaccine in reducing the severity of the infection. Likewise, the disease-induced death rate *δ*_2_ is lower for the vaccinated population. Individuals in the *I*_*V*_ and *A*_*V*_ classes also move to the *R* class upon recovery from the disease at a rate *γ*.

We will denote by *N* (*t*) the total population at time *t*, which is given by

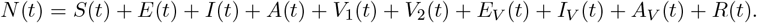

Hence, our model is described by the following system of differential equations:

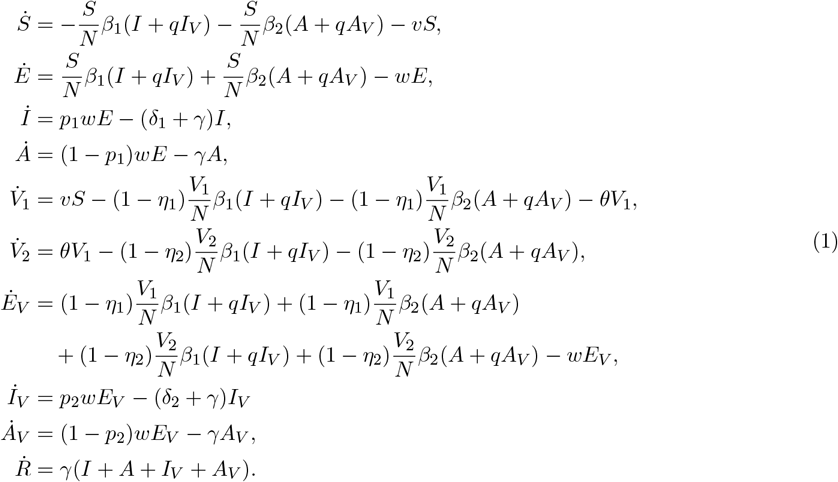

We define an additional variable *D*(*t*) that denotes the number of people deceased due to COVID-19, which is governed by the equation

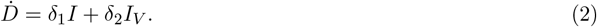

The flow diagram of the model can be seen in Figure 1. The list of parameters and their interpretation is as follows:

**Figure 1:**
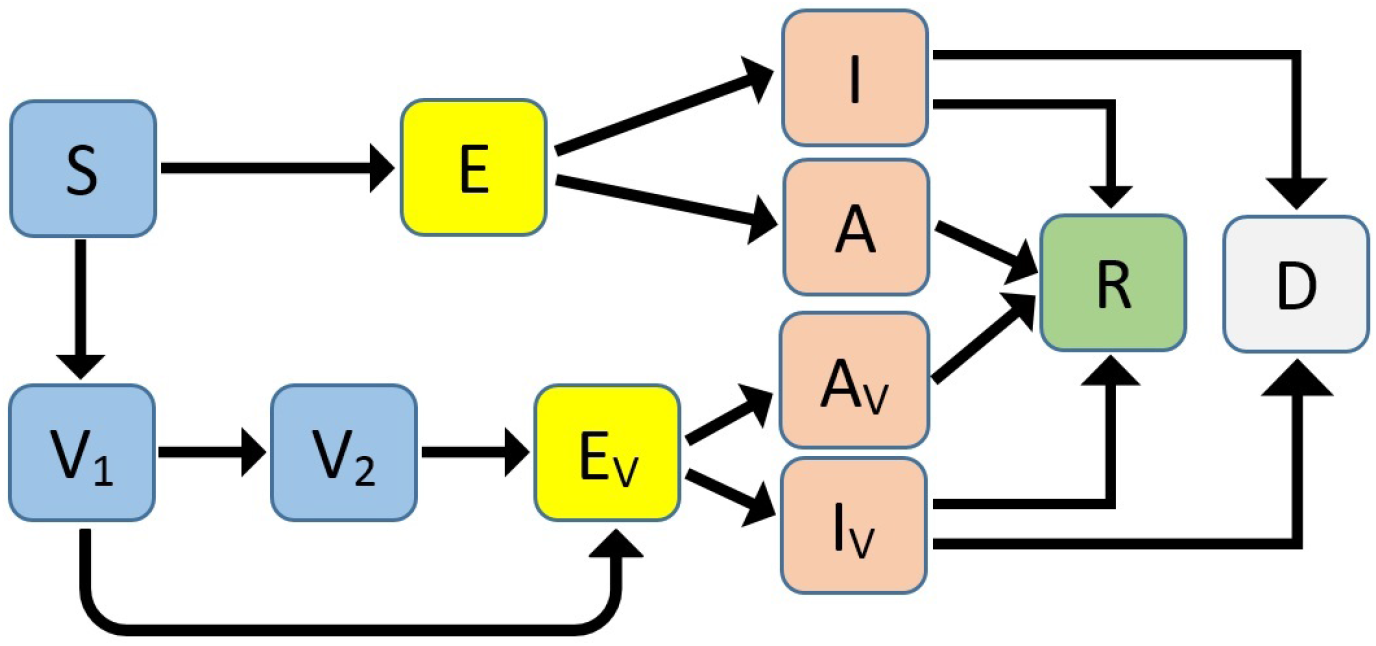
Flow diagram of the model with vaccination.

- *β*_1_ > 0: transmission rate by contact with symptomatic infectious individuals.
- *β*_2_ > 0: transmission rate by contact with asymptomatic infectious individuals.
- *q* ∈ [0, 1]: relative infectivity of individuals that become infected after vaccination.
- *v* ≥ 0: vaccination rate.
- 1*/θ*: time between the application of the first and the second dose of the vaccine.
- *η*_1_ ∈ [0, 1]: efficacy rate of the vaccine after the first dose.
- *η*_2_ ∈ [0, 1]: efficacy rate of the vaccine after the second dose.
- 1*/w*: length of the latent period.
- *p*_1_ ∈ [0, 1]: proportion of infectious unvaccinated individuals that show symptoms of the disease.
- *p*_2_ ∈ [0, 1]: proportion of infectious vaccinated individuals that show symptoms of the disease.
- *δ*_1_ ≥ 0: death rate of infectious unvaccinated individuals with symptoms.
- *δ*_2_ ≥ 0: death rate of infectious vaccinated individuals with symptoms.
- *γ >* 0: recovery rate.

## 3 Theoretical analysis

In this section, we will derive some theoretical results for model (1). Since we are mainly interested in the long term behaviour of the epidemic, we will focus on the case when the vaccination campaign is over and hence the vaccination rate *v* is zero, but the vaccinated subpopulations (*V*_1_, *V*_2_, *E*_*V*_, *I*_*V*_, *A*_*V*_) may have positive initial values.

First, we will determine the disease-free equilibria of the model. It is easy to notice that system (1) does not have any equilibria when *v* is positive. On the other hand, when *v* is zero, model (1) has a continuum of disease-free equilibria (DFE), given by

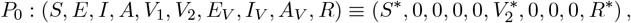

where *S*^*\**^ ≥ 0, 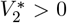 and *R*^*\**^ > 0. These equilibria represent the scenario when the anti-COVID vaccination programme has ended, and a certain number of individuals 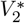 has been vaccinated to achieve herd immunity in the population. We will compute the *control reproduction number* ℛ_*c*_ of the model based on this expression for the DFE.

Using the notation in [29], we determine the matrix of new infections *F* and the transition matrix *V*, considering only the infected compartments (*E, I, A, E*_*V*_, *I*_*V*_ and *A*_*V*_). First, we define

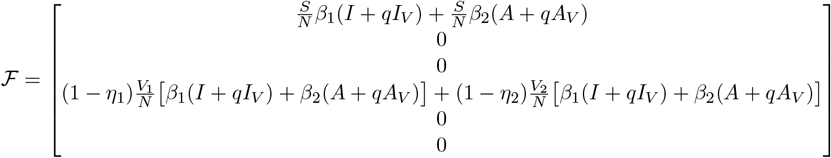

and

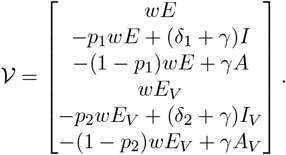

Then, the derivative of ℱ at a disease-free equilibrium *P*_0_ is

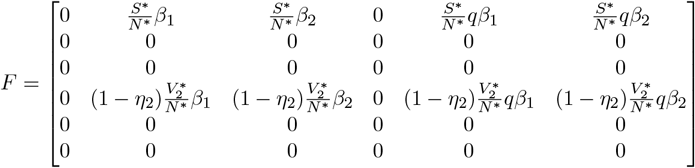

where 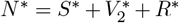 denotes the total population at the equilibrium. The derivative of 𝒱 evaluated at *P*_0_ is

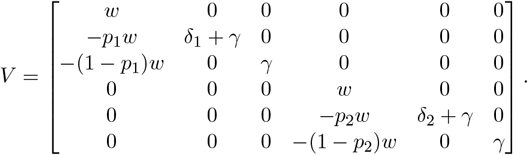

It follows that

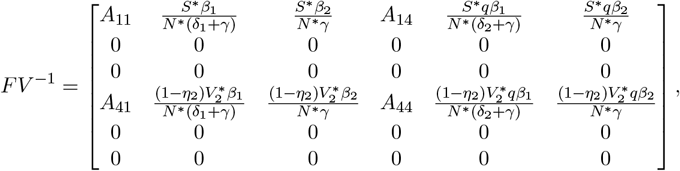

where

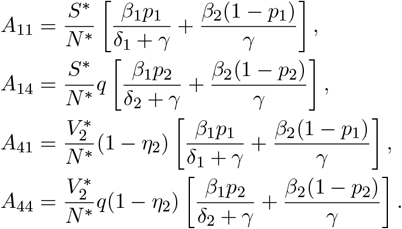

The control reproduction number ℛ_*c*_ of model (1) is given by ℛ_*c*_ = *ρ FV* ^*−*1^, where *ρ* denotes the spectral radius. Hence,

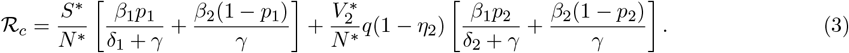

The quantity R_*c*_ measures the average number of new COVID-19 cases generated by a typical infectious individual introduced into a population where a fraction 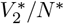 has been fully vaccinated using a vaccine with efficacy *η*_2_.

According to [29, Theorem 2], we can obtain the following result about the control reproduction number.

### Theorem 1.

*The continuum of disease-free equilibria P*_0_ *of system* (1) *with v* = 0 *is locally asymptotically stable if* ℛ_*c*_ < 1, *and it is unstable if* ℛ_*c*_ > 1.

The epidemiological interpretation of Theorem 1 is that, if ℛ_*c*_ < 1, then a small perturbation from a disease-free equilibrium will not generate an epidemic outbreak. On the other hand, if ℛ_*c*_ > 1, the epidemic curve will initially show an exponential growth, then reach a peak and start to decrease until becoming extinct.

The following theorem gives a sufficient condition for the global stability of the disease-free equilibria.

### Theorem 2.

*Suppose that*

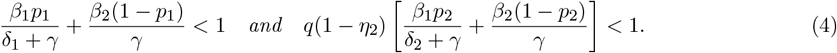

*Then, the continuum of disease-free equilibria P*_0_ *of system* (1) *with v* = 0 *is globally asymptotically stable*.

*Proof*. Consider the following Lyapunov function:

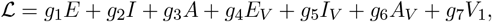

where

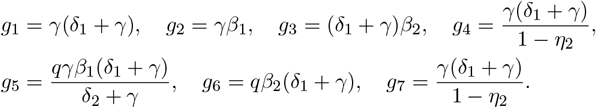

The time derivative of ℒ evaluated at the solutions of system (1) with *v* = 0 is given by

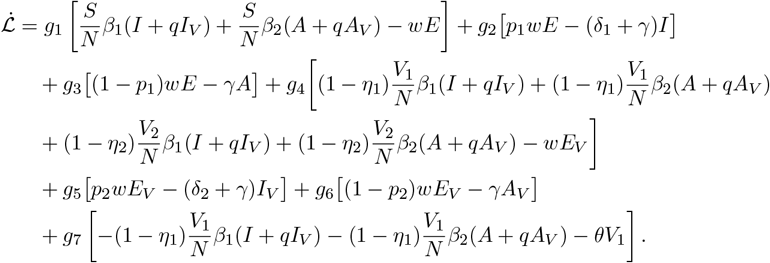

After cancelling terms and simplifying, we obtain

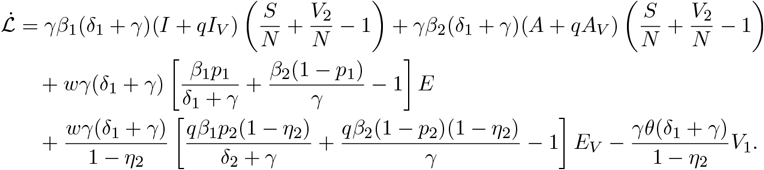

Since *S*(*t*) + *V*_2_(*t*) ≤ *N* (*t*) for all *t*, we have 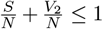. Combining this with the hypothesis (4), we can see that 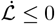, and 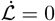 if and only if *E*(*t*) = 0 and *E*_2_(*t*) = 0. Substituting *E*(*t*) = 0 and *E*_2_(*t*) = 0 in system (1) with *v* = 0 shows that 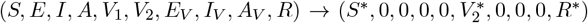 as *t* → ∞. Hence, the largest positively invariant set where 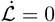 is the continuum of disease-free equilibria. Therefore, by LaSalle’s invariance principle, we conclude that *P*_0_ is globally asymptotically stable.

## 4 Numerical simulations

In this section, we perform some numerical simulations for model (1) to provide estimates for the evolution of the COVID-19 outbreak in Mexico.

### 4.1 Data fitting and estimation of parameters

We used cumulative data provided by the Johns Hopkins University repository [30] to fit the parameters of model (1) in the absence of vaccination. We considered the data for reported COVID-19 infections, deaths and recovered cases during the period from 12 November 2020 to 24 December 2020, which is before the vaccination programme in Mexico began.

For this part, we considered system (1) with *v* = 0 and the vaccinated subpopulations *V*_1_, *V*_2_, *E*_*V*_, *A*_*V*_ and *I*_*V*_ equal to zero. We regarded as fixed parameters *w* = 0.25, which corresponds to a latent period of 4 days [31], and a proportion *p*_1_ = 0.12 of symptomatic infections [28]. The set of differential equations was solved using Matlab 2016b with the ode45 solver. The values for *β*_1_, *β*_2_, *δ*_1_ and *γ* were estimated by minimizing the Sum of Squared Errors (SSE), which is calculated as follows. For a given vector of parameters **x**, we compute numerically the *I*(*t*) and *D*(*t*) components of the solutions for our model, as well as the estimated number of people recovered from symptomatic infections *R*_*I*_(*t*) and the cumulative number of symptomatic infected cases *C*(*t*), defined by

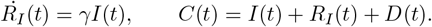

Then the Sum of Squared Errors is given by

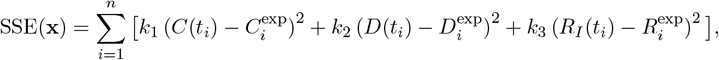

where 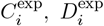 and 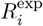 denote the experimental data for cumulative infections, deaths and recoveries, respectively, reported for day *t*_*i*_ (*i* = 1, …, *n*), while *k*_1_, *k*_2_ and *k*_3_ are coefficients used to compensate the order of magnitude for the data. In our simulations, we used *k*_1_ = 20, *k*_2_ = 10 and *k*_3_ = 1. The global minimum of the SSE function was obtained by applying three consecutive searches: a gradient-based method, a gradient-free algorithm and, again, a gradient-based method.

The best fit values for *β*_1_, *β*_2_, *δ*_1_ and *γ* are shown in Table 1. Figure 2 depicts a comparison between the model solutions and the observed cumulative COVID-19 data before the vaccination period.

**Table 1:** Baseline values for the parameters used in the simulations.

| Parameter | Description | Value | Source |
| --- | --- | --- | --- |
| $\beta_1$ | Transmission rate by contact with $I$ and $I_V$ classes | $0.2 \text{ day}^{-1}$ | Fitted to data |
| $\beta_2$ | Transmission rate by contact with $A$ and $A_V$ classes | $0.0330 \text{ day}^{-1}$ | Fitted to data |
| $q$ | Relative infectivity of individuals in $I_V$ and $A_V$ classes | 0.52 | [32] |
| $v$ | Vaccination rate | Variable | Assumed |
| $\theta$ | Application rate of second doses | $1/70 \text{ day}^{-1}$ | Assumed |
| $\eta_1$ | Vaccine efficacy rate after one dose | 0.463 | [33] |
| $\eta_2$ | Vaccine efficacy rate after two doses | 0.557 | [33] |
| $w$ | Transfer rate from exposed to infectious | $0.25 \text{ day}^{-1}$ | [31] |
| $p_1$ | Portion of people in $E$ class that develop symptoms | 0.12 | [28] |
| $p_2$ | Portion of people in $E_V$ class that develop symptoms | 0.089 | Estimated |
| $\delta_1$ | Death rate of $I$ class | $3.2135 \times 10^{-3} \text{ day}^{-1}$ | Fitted to data |
| $\delta_2$ | Death rate of $I_V$ class | 0 | Assumed |
| $\gamma$ | Recovery rate of infectious individuals | $3.6987 \times 10^{-2} \text{ day}^{-1}$ | Fitted to data |

**Figure 2:**
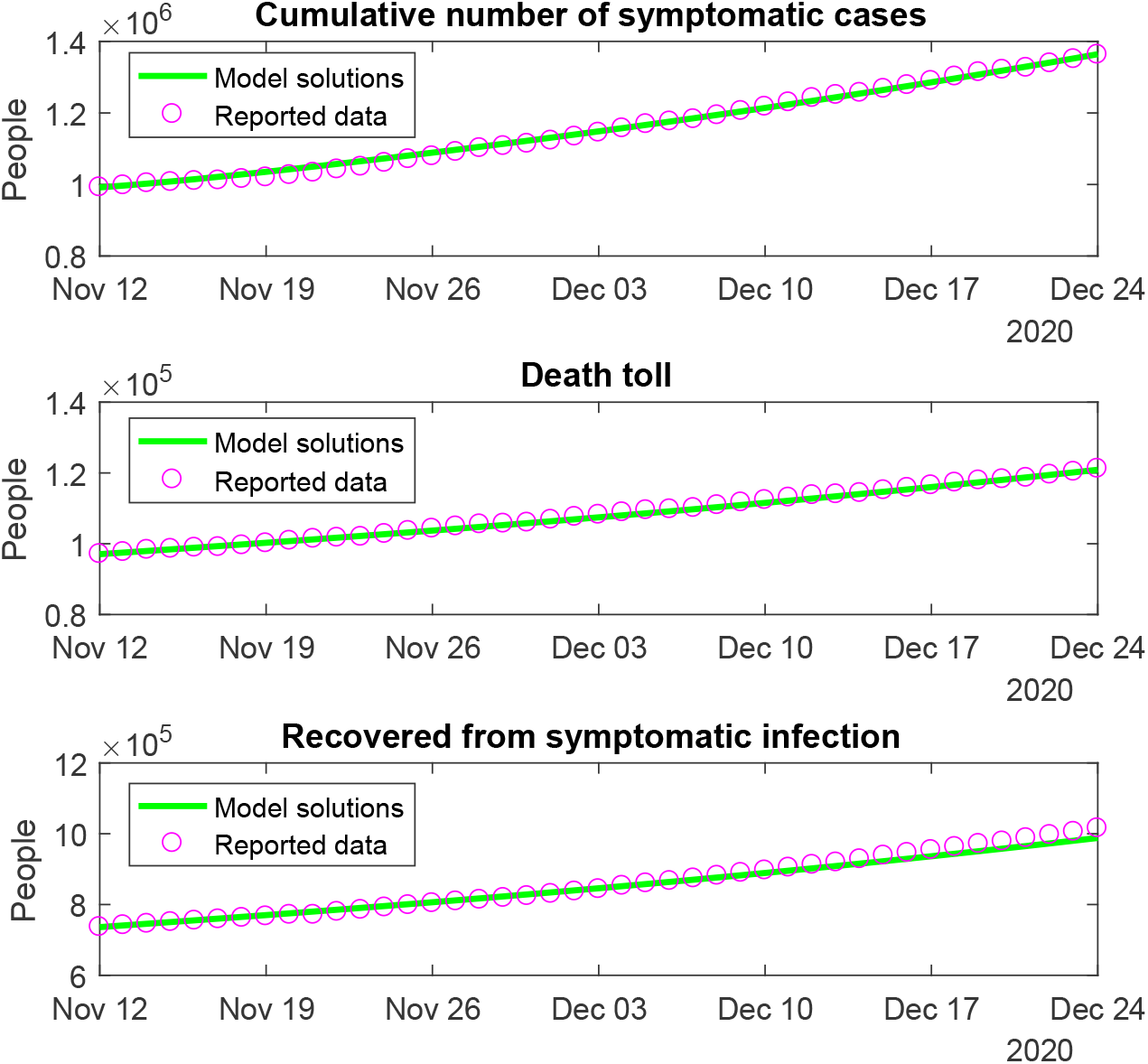
Reported cumulative number of symptomatic cases, COVID-19 deaths and recovered cases in Mexico for the pre-vaccination period, and simulations using model (1) with the parameters in Table 1 and *v* = 0.

### 4.2 Simulations for the model with vaccination

We will now simulate the solutions to model (1) to assess the impact of the vaccination campaign that started in Mexico in December 2020 to combat the COVID-19 pandemic.

As of August 2021, seven COVID-19 vaccines have received Emergency Use Authorization for their deployment in Mexico:

- BNT162b2 (Pfizer–BioNTech),
- AZD1222 (Oxford–AstraZeneca),
- Sputnik V (Gamaleya Institute),
- CoronaVac (Sinovac),
- BBV152 (Covaxin),
- Ad5-nCoV (CanSino), and
- Ad26.COV2-S (Johnson & Johnson).

The first five of these vaccines require two doses, while CanSino and Johnson & Johnson are single-dose vaccines [34].

Efficacy estimates for each vaccine based on data from clinical trials are subject to change with the emergence of new analyses. An interim analysis for the Oxford–AstraZeneca vaccine [33] estimated an efficacy against infection (symptomatic or asymptomatic) of 46.3% (31.8%–57.8%), considering people who had a nucleic acid amplification test (NAAT)-positive swab more than 21 days after a single dose, and 55.7% (41.1%–66.7%) for people who tested positive more than 14 days after a second dose of the vaccine. However, a more recent study [35] estimated an efficacy of 63.9% (46.0%–76.9%) after one dose and 59.9% (35.8%–75.0%) after two standard doses given 12 or more weeks apart.

Due to longer dose intervals being associated with greater efficacy against symptomatic infection, the WHO has recommended to administer the Oxford–AstraZeneca vaccine with an interval of 8 to 12 weeks between first and second doses [36]. Based on the above, we will assume in our simulations an average length of 1*/θ* = 70 days for the inter-dose period, and we will use *η*_1_ = 0.463 and *η*_2_ = 0.557 as baseline values for the efficacy parameters. Furthermore, we assume a reduction of 48% in the infectivity of individuals becoming infected after being vaccinated (i.e., *q* = 0.52), following the estimations in [32].

For computing the proportion of infectious vaccinated individuals that show symptoms of the disease (*p*_2_), we follow [33], who reported 37 cases of symptomatic COVID-19 disease out of a total of 68 NAAT-positive swabs in the group of people vaccinated with AZD1222, and 112 symptomatic cases out of 153 NAAT-positive cases in the control group. This yields a reduction from 0.732 to 0.544 in the symptomatic proportion after vaccination. Since we have chosen *p*_1_ = 0.12, we will take *p*_2_ = 0.089 so that *p*_1_ : *p*_2_ = 0.732 : 0.544. Furthermore, we assume that the death rate *δ*_2_ of infectious vaccinated people is zero since it is widely accepted that current anti-COVID vaccines provide full protection against severe infections.

We used the daily data on COVID-19 vaccinations in Mexico obtained from [37] to estimate the value of the vaccination rate *v* over nine different date ranges, as shown in Table 2. We plot in Figure 3 a comparison of the reported number of vaccinated people and the simulations obtained with model (1) for the period 24 December 2020 – 27 July 2021. In these graphs, we considered the total population of Mexico as 127 090 000 people.

**Table 2:** Estimated values for the vaccination rate *v*.

| Date | Value ($\text{day}^{-1}$) |
| --- | --- |
| 24 Dec 2020 – 11 Jan 2021 | $4.0 \times 10^{-5}$ |
| 12 Jan 2021 – 15 Jan 2021 | $7.9 \times 10^{-4}$ |
| 16 Jan 2021 – 14 Feb 2021 | $6.0 \times 10^{-5}$ |
| 15 Feb 2021 – 7 Mar 2021 | $7.3 \times 10^{-4}$ |
| 8 Mar 2021 – 14 Mar 2021 | 0.0021 |
| 15 Mar 2021 – 31 Mar 2021 | 0.0017 |
| 1 Apr 2021 – 15 Apr 2021 | 0.0035 |
| 16 Apr 2021 – 15 May 2021 | 0.0017 |
| 16 May 2021 – 27 Jul 2021 | 0.0060 |

**Figure 3:**
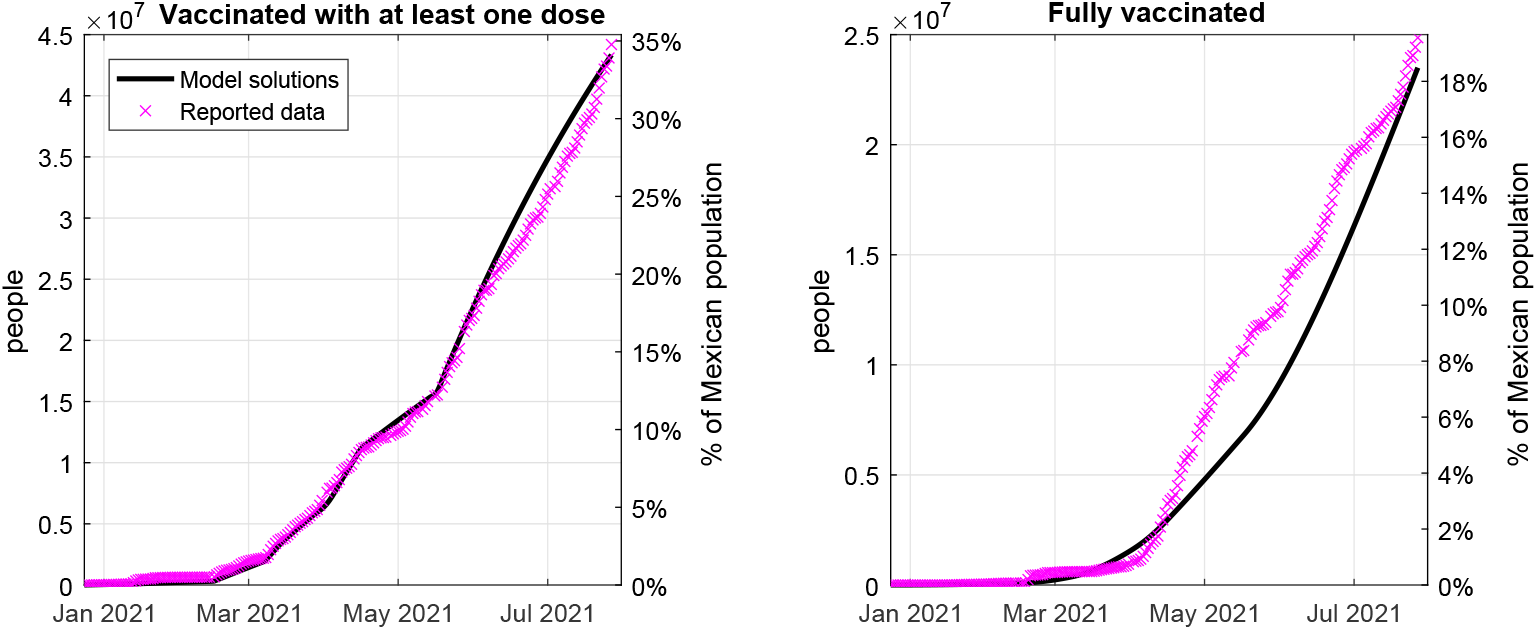
COVID-19 vaccination coverage in Mexico from 24 December 2020 to 27 July 2021. X represents real data, continuous lines represent model simulations using the vaccination rate in Table 2.

In order to obtain long-term projections for the vaccination coverage in Mexico, we simulated two different scenarios. First, we assumed that the vaccination rate is kept constant at its baseline value on 27 July 2021 (0.6% of susceptible population per day, which equals roughly 294 000 first doses per day). Second, we assumed that the vaccination rate increases to twice its baseline value starting on September 2021. Figure 4 shows that, if vaccines continue to be delivered at their baseline rate, the vaccination coverage with at least one dose will have reached 70% of Mexican population by January 2023. On the other hand, if the vaccination rate is doubled, the same coverage with at least one dose could be achieved by May 2022, and 70% of the Mexican population could be fully vaccinated by September 2022.

**Figure 4:**
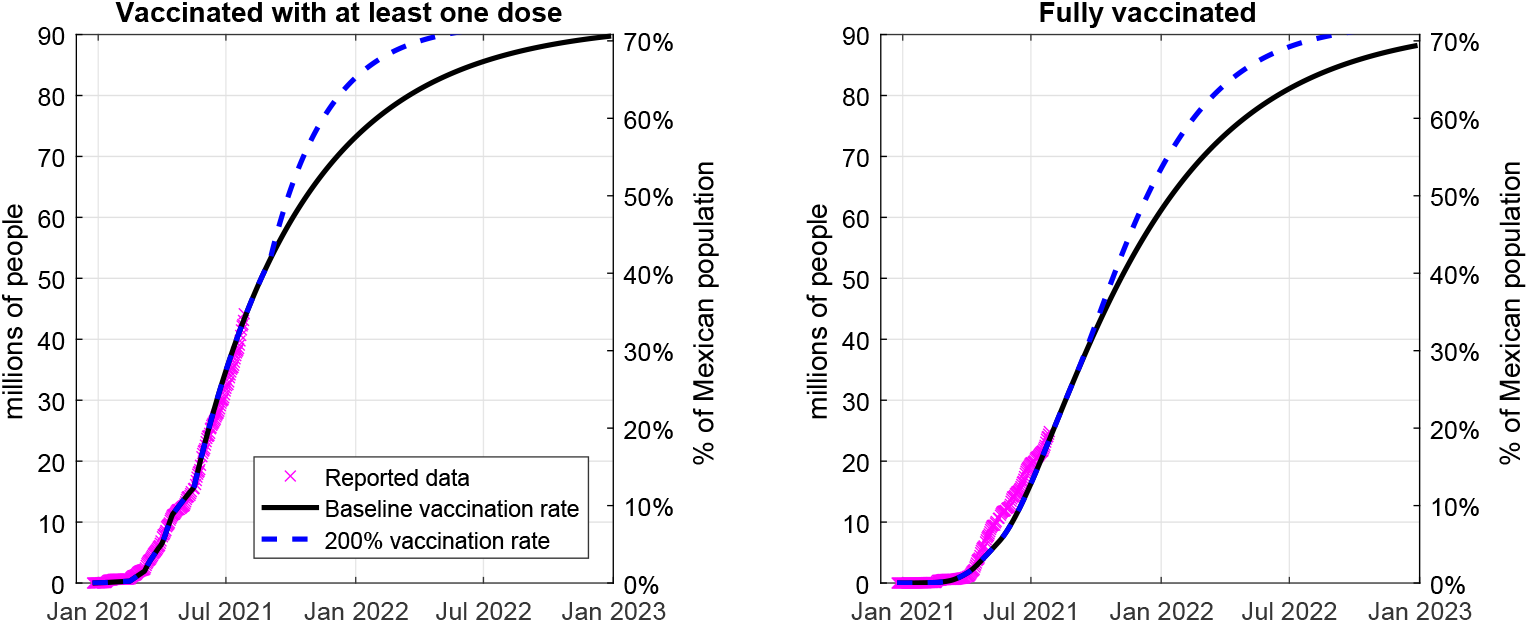
Long-term projections of COVID-19 vaccination coverage in Mexico. X represents real data, continuous lines represent simulations using the baseline vaccination rate, and dashed lines represent simulations using 200% vaccination rate from September 2021 onwards.

#### 4.2.1 Assessing the effect of vaccination and different transmission rates

We will next compute the solutions of model (1) to simulate the evolution of the pandemic in Mexico as the vaccination campaign takes place. We consider the initial date for simulations as 24 December 2020. Based on the results obtained in Subsection 4.1, we use the initial conditions

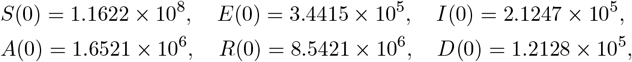

and *V*_1_(0) = *V*_2_(0) = *E*_*V*_ (0) = *I*_*V*_ (0) = *A*_*V*_ (0) = 0. In these subsection, we will consider different values for the transmission rates *β*_1_ and *β*_2_ to account for the possibility that the number of infectious contacts between people may increase or decrease due to resumption of economical activities, compliance with social/physical distancing, wearing of face masks, etc. Hence, we consider three cases: when *β*_1_ and *β*_2_ are kept with the values in Table 1, when both of them decrease to an 80% of these values, and when they increase to a 120%. The values for other parameters are fixed as in Tables 1 and 2.

Figure 5 depicts the time evolution of the number of infectious COVID-19 cases with symptoms (*I*(*t*)+*I*_*V*_ (*t*)) and the death toll (*D*(*t*)) for each of the above cases. In each graph, we have plotted the solutions assuming the baseline vaccination rate and the 200% vaccination rate, as well as a counterfactual case with no vaccination.

**Figure 5:**
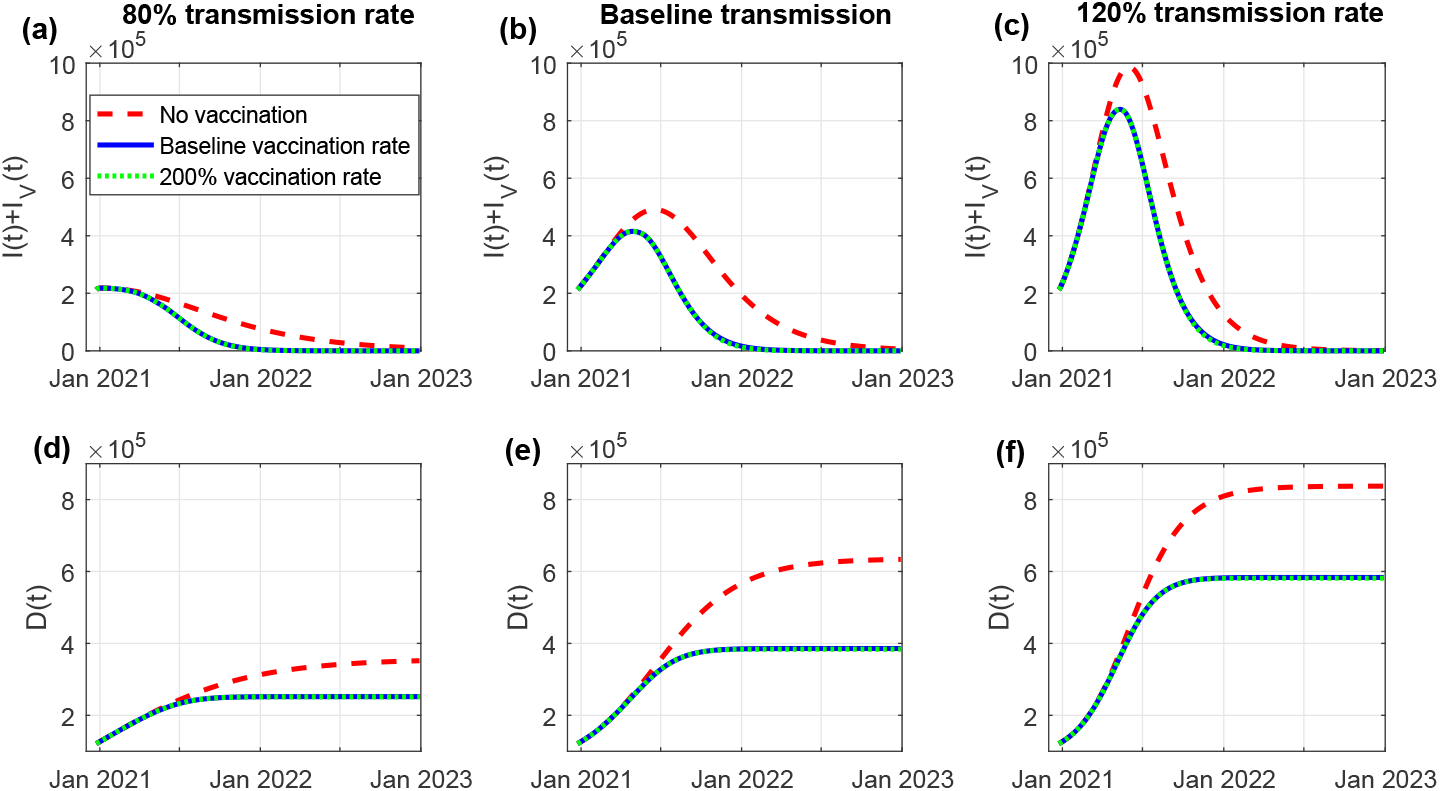
Simulations of model (1) using different values for the transmission rates. (a) and (d): 80% transmission rate. (b) and (e): baseline transmission rate. (c) and (f): 120% transmission rate. Top row: number of active symptomatic infectious cases. Bottom row: Cumulative number of deaths.

Figure 5(a) shows that, in the case of low transmission rate, the number of active cases would start to decrease in the early months of 2021, and the epidemic would be almost extinguished by January 2022. In the cases with higher transmission rate (Figures 5(b) and (c)), the epidemic curve would reach its peak around May 2021, and the number of active symptomatic cases would be less than 1000 by May 2022. Figures 5(d)–(f) show that the cumulative number of deaths would be around 250 000 for low transmission, 390 000 for baseline transmission, and 580 000 for high transmission rates.

We can also see that an increase in the vaccination rate to double its baseline value does not result in a considerable change in the number of infections or deaths, although the vaccination scenarios result in around 250 000 less deaths compared with the case with no vaccination. On the other hand, comparing Figures 5(d) and (e) shows that more than 130 000 deaths can be avoided by reducing the transmission rate to 80%, while a 20% increase in the transmission rate would result in almost 200 000 additional deaths (Figure 5(f)). This suggests that decreasing the number of infectious contacts by complying with preventive measures is more effective than simply accelerating the deployment of vaccines.

#### 4.2.2 Assessing the effect of different vaccine efficacy rates

Given that there is still uncertainty regarding the efficacy of anti-COVID vaccines against infection, including asymptomatic cases, we will also simulate the solutions of model (1) using different values for the parameters *η*_1_ and *η*_2_.

Figure 6 shows the number of active infectious cases with symptoms (*I*(*t*) + *I*_*V*_ (*t*)) and without symptoms (*A*(*t*) + *A*_*V*_ (*t*)), as well as the death toll (*D*(*t*)), using different efficacy rates: in addition to the baseline case (*η*_1_ = 0.463, *η*_2_ = 0.557), we include a case with lower efficacy (*η*_1_ = 0.4, *η*_2_ = 0.45) and a case with higher efficacy (*η*_1_ = 0.6, *η*_2_ = 0.65). Here, we have plotted all solutions using the baseline vaccination rate. The simulations show that lower efficacy results in an additional 7 429 symptomatic cases and 79 624 asymptomatic cases at the peak of the infection curve, compared with the case with higher efficacy. However, this does not significantly affect the time when the peak occurs. Moreover, lower efficacy also results in 4 953 additional deaths.

**Figure 6:**
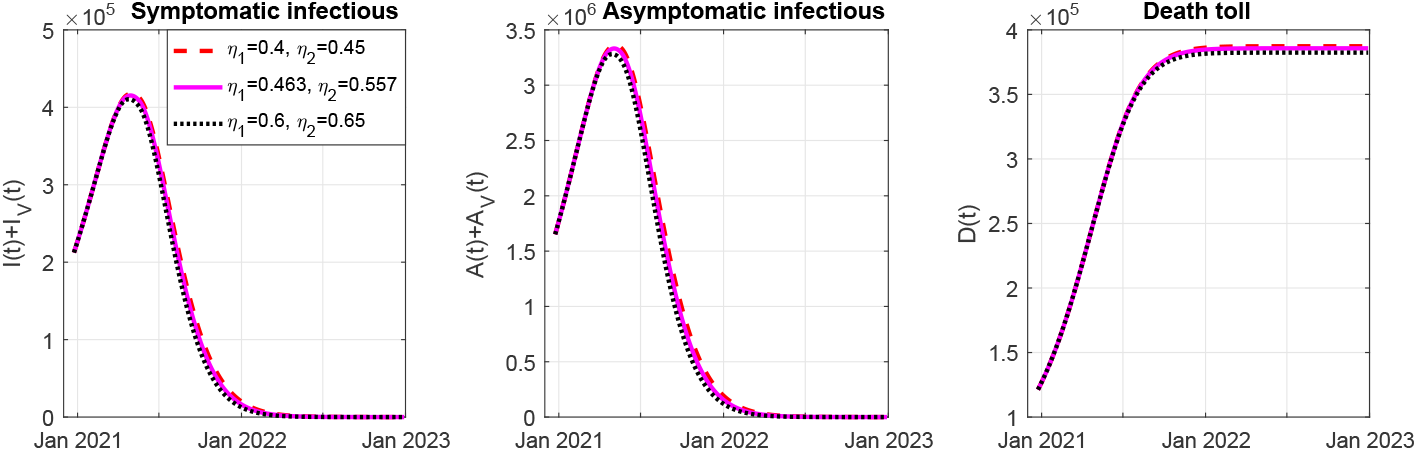
Simulations of model (1) using different values for the vaccine efficacy rates. Left panel: number of active symptomatic infectious cases. Central panel: number of active asymptomatic infectious cases. Right panel: cumulative death toll.

### 4.3 Impact of vaccination coverage on the control reproduction number

Next, we will study how the control reproduction number ℛ_*c*_ is affected by some of the model parameters.

By equation (3), we know that ℛ_*c*_ does not only depend on the parameters of system (1), but also on the final proportions of unvaccinated susceptible people (*S*^*\**^*/N*^*\**^) and fully vaccinated people 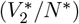 at the time when vaccines are no longer being deployed to the population.

We recall that a disease-free equilibrium takes the form 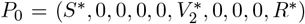, where the total population is 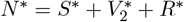. If we define

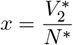

as the proportion of fully vaccinated people and

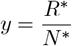

as the proportion of people recovered from COVID-19, we can rewrite the expression for the control reproduction number as

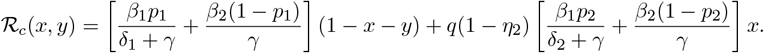

Figure 7 depicts the value of ℛ_*c*_ as function of the proportions *x* and *y*, using several values for the transmission rates and efficacy after the second vaccine dose. Other parameter values were taken as in Table 1. We can see that an increase in either *x* or *y* contributes to reducing the reproduction number, and therefore, is helpful towards achieving herd immunity.

**Figure 7:**
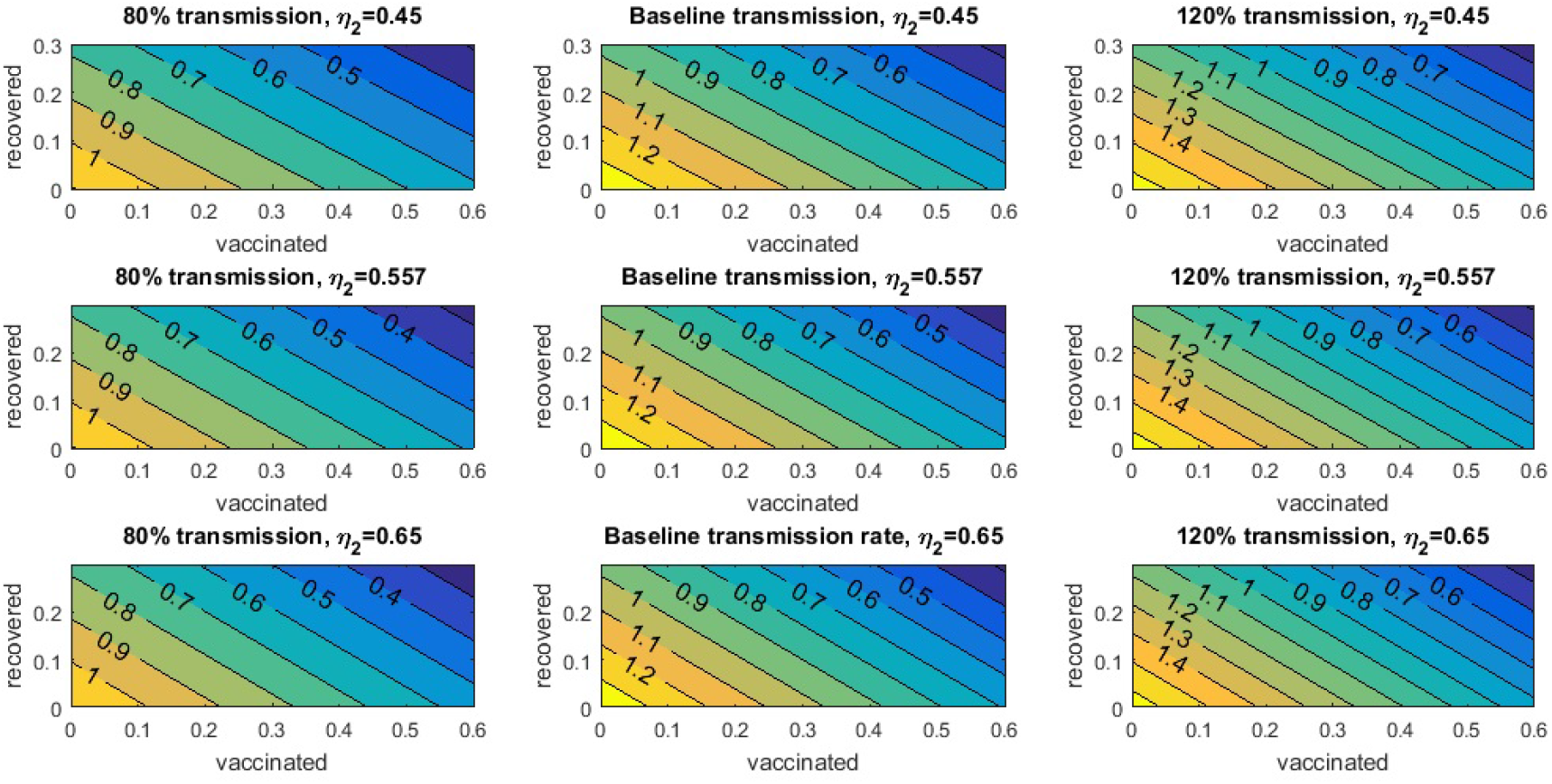
Value of the control reproduction number as function of the proportion of fully vaccinated individuals (horizontal axis) and recovered individuals (vertical axis).

Herd immunity occurs when a large portion of the population has become immune to the disease due to vaccination or natural recovery, which makes spread of the disease difficult. Thus, the minimal level of vaccination coverage that is required to achieve herd immunity (that is, making ℛ_*c*_ < 1) will also depend on the percentage of the population that has been infected and then successfully recovered. Comparing the different panels of Figure 7, we can see that increasing the vaccine efficacy *η*_2_ reduces the vaccination coverage needed to make ℛ_*c*_ < 1 for a fixed proportion of recovered people. However, this reduction is small compared to the effect gained by decreasing the transmission rate. For example, when *η*_2_ = 0.65 and the recovered population is close to zero, it is necessary to vaccinate 46% of population to obtain ℛ_*c*_ = 1 in the case of 120% transmission rate, 33% in the case of baseline transmission rate, and only 11% of population in the case of 80% transmission rate (bottom row of Figure 7).

## 5 Conclusion

In this work, we studied a model for COVID-19 with vaccination. Our work was based on the SEIARD model proposed in [28], which included an exposed (latent) compartment and different transmission rates for the symptomatic and asymptomatic infectious individuals; we extended this model by incorporating vaccinated compartments and considering a two-dose vaccination regime. Although several COVID-19 models with vaccination have been proposed in the literature, most studies have focused only on carrying out numerical simulations, while our work shed some light on the theoretical properties of model (1).

We showed that our model has multiple disease-free equilibria and computed the control reproduction number ℛ_*c*_ using the next-generation matrix method. We established that the set of disease-free equilibria is locally asymptotically stable when ℛ_*c*_ < 1 and unstable when ℛ_*c*_ > 1. Furthermore, we determined a condition that guarantees the global asymptotic stability of the DFE.

We performed a numerical simulation on our model using repository data on the outbreak of COVID-19 in Mexico and the daily data on COVID-19 vaccinations to estimate the value of the vaccination rate over nine different date ranges. We used the efficacy estimates based on data from clinical trials of the Oxford–AstraZeneca vaccine, which is the one that is being more widely distributed in Mexico at the time of this writing. We remark that, in this article, we considered *vaccine efficacy* in the sense of protection against COVID-19 infection (symptomatic or asymptomatic), while other works consider efficacy as protection against symptomatic infection only.

In order to obtain long-term projections for the vaccination coverage, we simulated two different scenarios. First, we assumed that the vaccination rate is kept constant by vaccinating the same proportion of susceptible individuals per day, and secondly, we assumed that the vaccination rate increases to twice its baseline value from September 2021 onwards. Our study showed that if vaccines continue to be delivered at their baseline rate, by January 2023 the first dose will be applied to 90 million people, which represents roughly the total Mexican population over age 18. On the other hand, if the vaccination rate is doubled, the total adult population could be partially vaccinated by May 2022 and fully vaccinated by September 2022. In the case of low transmission rate, the number of active cases would start to decrease in the early months of 2021, and the epidemic would be almost eradicated in early 2022, while in the cases with medium to high transmission rate the epidemic curve would reach its peak around May 2021 and would be close to zero by mid-2022.

Our simulations show that keeping a low transmission rate (by wearing face masks, complying with social/physical distancing, etc.) is the most effective method to reduce the death toll. For example, reducing the transmission rate to 80% its baseline value results in 130 000 less deaths, while doubling the vaccination rate does not yield a significant reduction in death toll. Also, decreasing the transmission rate is more effective to reduce the control reproduction number and achieve herd immunity than deploying vaccines with higher efficacy rates.

Our model has certain limitations that could affect the results presented in this study. Firstly, our model assumes that the time between first and second doses and the efficacy rate are the same for all vaccines applied in the population; in reality, several vaccines (including single-dose vaccines) may be applied simultaneously in the same country, which would yield different parameter values for each of them. Secondly, we assumed that the protection against COVID-19 provided by vaccines does not wane over time since research about the immunity waning rate is still ongoing. Thirdly, we assume that the contact rate remains constant throughout the simulations, when it could actually change at different times due to lifting of restrictions and lockdown in the population. Lastly, the emergence of different variants of SARS-CoV-2 could be taken into account to consider the increased transmissibility and possible reduction of vaccine efficacy against these variants. These shortcomings will be addressed in future studies.

## Data Availability

The data used in this manuscript were publicly available data retrieved from Johns Hopkins University Repository and Our World in Data.

https://github.com/CSSEGISandData/COVID-19

https://github.com/owid/covid-19-data/tree/master/public/data/vaccinations

